# Oscillatory dynamics in infectivity and death rates of COVID-19

**DOI:** 10.1101/2020.05.19.20107474

**Authors:** Tomáš Pavlíček, Pavel Rehak, Petr Král

**Author notes:** Addresses for Correspondence.

## Abstract

The analysis of systematically collected data for COVID-19 infectivity and death rates has revealed in many countries around the world a typical oscillatory pattern with a 7-days (circaseptan) period. Additionally, in some countries the 3.5-days (hemicircaseptan) and 14-days periodicities have been also observed. Interestingly, the 7-days infectivity and death rates oscillations are almost in phase, showing local maxima on Thursdays/Fridays and local minima on Sundays/Mondays. These observations are in stark contrast with a known pattern, correlating the death rate with the reduced medical staff in hospitals on the weekends. One possible hypothesis addressing these observations is that they reflect a gradually increasing stress with the progressing week, which can trigger the maximal death rates observed on Thursdays/Fridays. Moreover, assuming the weekends provide the likely time for new infections, the maximum number of new cases might fall again on Thursdays/Fridays. These observations deserve further study to provide better understanding of the COVID-19 dynamics.

## Introduction

The current worldwide pandemic caused by the SARS-HCoV-2 coronavirus has led within several months to millions of infected individuals and hundreds of thousands of fatalities, while the economies of most countries have been largely put on stand style. Therefore, it is urgent to understand the dynamics of coronaviruses since their repeated straying into the human population could provoke an existential crisis. As a recent report warns^1^, dramatic interventions by the individual governments to locally reduce the effective reproductive number (*R_t_*) by a lockout might not substantially change the long-term, total number of infections, and possible fatalities, if the human behavior returns to normal before a vaccine is available^2^. Moreover, with the onset of the winter season in the Southern Hemisphere, the COVID-19 infections seem to largely follow an exponentially increasing trend, which is expected to reappear as a second wave in the Northern Hemisphere.

Subfamily *Coronavirinae^3^* house the following four genera: *Alphacoronavirus* (contains many animal and two human coronaviruses, HCoV-229E, and HCoV-NL63), *Betacoronavirus* (contains mouse hepatitis virus, MHV, and the following human viruses: HCoV-OC43, HCoV-HKU1, Middle East Respiratory Syndrome Coronavirus, MERS-HCoV, Severe Acute Respiratory Syndrome Coronavirus SARS-HCoV, and SARS-HCoV-2), *Gamacoronavirus* and *Deltacoronavirus* (contains viruses from cetaceans, birds and pigs)^3^. As of today, at least seven coronaviruses are already circulating in the human population. However, SARS-HCoV-2 has a lot of peculiarities, probably caused by a strong and selective binding of its spike protein to human angiotensin-converting enzyme 2 (ACE2)^4^ One of the less obvious peculiarities is a periodic appearance of COVID-19 infectivity and death rates with a typical 7-days oscillatory pattern observed in numerous countries (https://www.worldometers.info/coronavirus/).

Chronobiology describes the mechanisms underlying chronomes, structures in time, which are found in individual organisms, in populations, and in the environment^5^. About 7-days (circaseptan) periodicity is frequently found among plants and animals including humans. As an example of the circaseptan periodicity, we can mention the following cases that might be triggered by internal or external factors^6^:

- the growth of tail and in oviposition in springtails^7^,
- the occurrence of cardiovascular events in humans^8^,
- the rejection of allografts in rats and humans^9^,
- the post-surgical swelling decreased after maxillo-facial surgery in humans^10^,
- the many meteorological and pollution variables^11^,
- the societal habits relating to the periods of rest and activity^6^.

In contrast to the circadian clock^12^, there isn’t yet any evidence about a molecular mechanism causing the circaseptan periodicity. Interestingly, the circaseptan periodicity in humans might gave rise to the 6-days working week followed by 1 day of the rest, observed around the world. In this weekly periodicity, COVID-19 emerged and possibly locked to the natural circaseptan periodicity, while magnifying it through the observed periodicity in infectivity and death rates. In certain countries, the weekly repeating maxima and minima of COVID-19 infectivity and related death rates are emerging at certain days with a probability close to certainty. It is particularly striking that the periodicities of COVID-19 infectivity and death rates are almost in phase, and the same pattern is separately followed by very different countries. Even though, these effects could be to some extent caused by periodic oscillations in human measuring and reporting of these events, we cannot exclude more profound reasons of these observations.

## Results

A Fourier transform was used to calculate the spectra associated with the dynamics of new cases and deaths per day of COVID-19, and to reveal possible periodicities present in the reported data in different countries. A detail analysis was only done in a small set of countries from Europe, Americas, and Asia, as well as from the whole World. We did not analyze the population in countries which reported nearly zero number of cases of the coronavirus infections and countries where the reported numbers of newly acquired cases and deaths could be less reliable.

### Europe

Italy was the first European country which was very early and severely hit by COVID-19. It did not have enough time to prepare and react adequately to the severity of the situation and operated under a large stress, while gradually strengthening the lock down rules. Figure 1 (top left) reveals a strong 7-days and a weak 3.5-days periodicities present in its infectivity, but much weaker oscillations in the death rate (top right). The minima of infectivity usually took part on Sundays and death rate on Mondays, while the maxima were about a half week shifted from the minima.

**Fig. 1.**
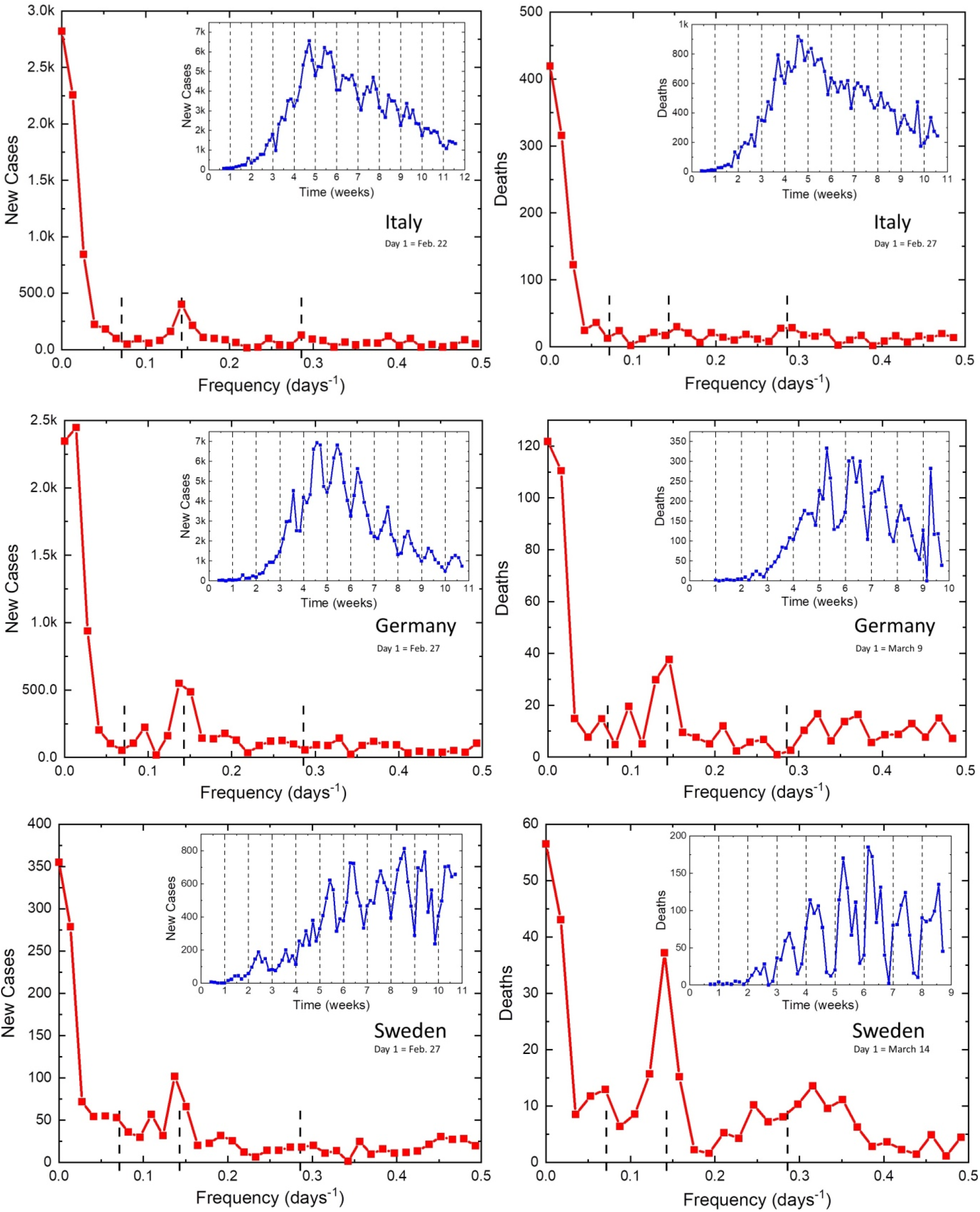
Daily new cases and deaths of COVID-19 reported in Italy (top), Germany (middle), and Sweden (bottom).

In contrast, Germany has developed COVID-19 infections later and did not implement restrictions for a relatively long time. Figure 1 (middle) reveals very strong oscillations with a 7-days period present in both studied parameters. These oscillations could reflect the free activity of the virus without limitations caused by a lock down. Even more relaxed approach was followed in Sweden, which was fully open for a long time. Figure 1 (bottom) reveals the strongest observed oscillations present in both parameters, especially in the death rate. Here, we can also clearly observe the 14-days and 3.5-days oscillations.

Other large European countries, such as Spain and France have been also severely hit by COVID-19, but the periodic oscillations in the observed parameters were not clearly enough seen, possibly due to more restricted and chaotic development of the situation. Other smaller countries have also shown medium oscillatory patterns. From the other large European countries, UK (Fig. 2 (top)) has relatively well developed 7-days and a weak 3.5-days periodicities, perhaps due to relatively relaxed lock down rules. However, these oscillations were absent in Poland (Fig. 2 (bottom)).

**Fig. 2.**
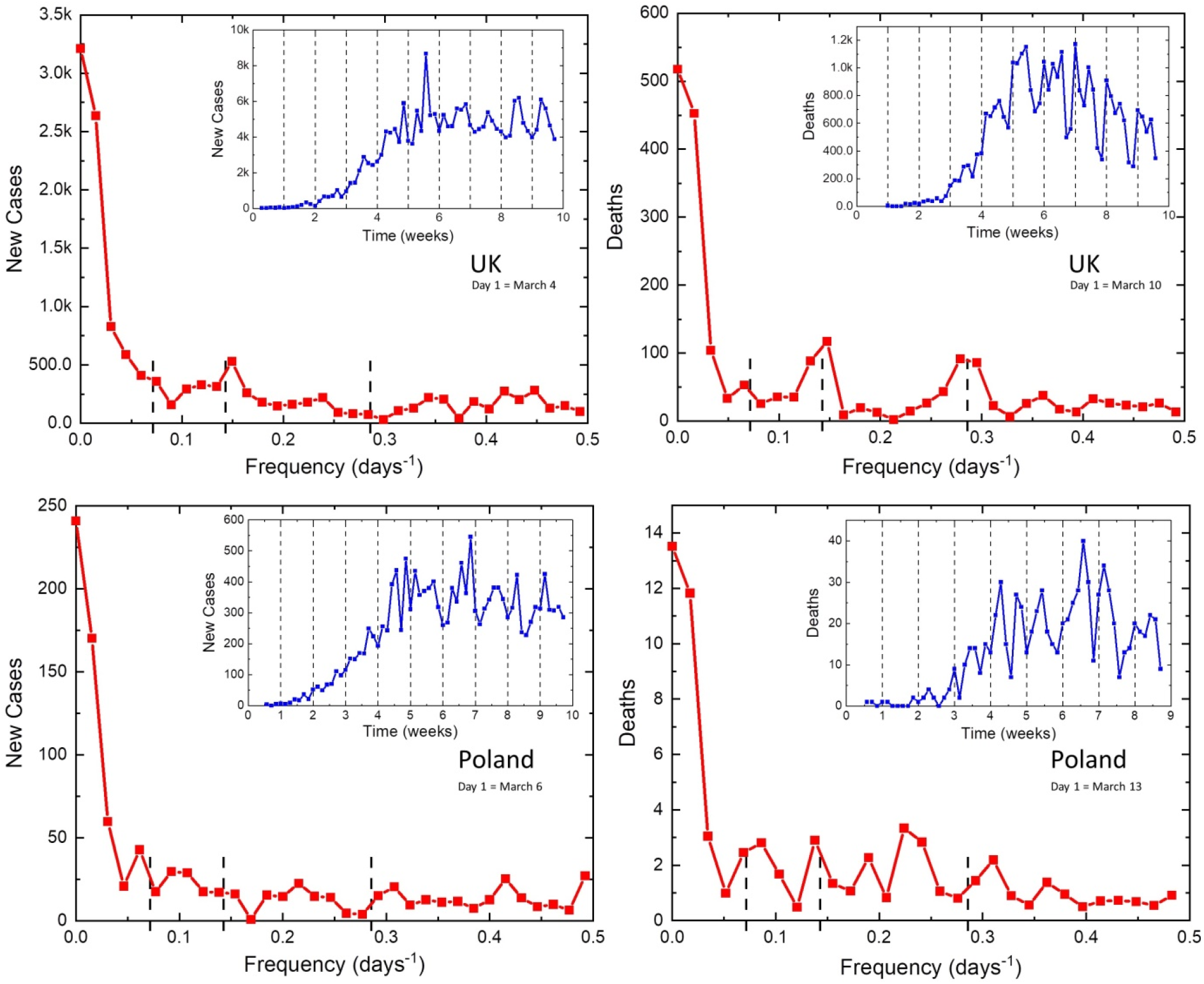
Daily new cases and deaths of COVID-19 reported in UK (top) and Poland (bottom).

### Americas

A similar pattern of the COVID-19 infection has developed in Americas. US (Fig. 3 (top)) initially seemed to follow the pattern like in Italy (NY). Later its oscillatory response turned out to be similar like in UK, where well developed 7-days and a weak 3.5-days periodicities were observed. In contrast, Mexico (Fig. 3 (middle)) and other countries in South America showed relatively strong oscillations with 14, 7, and 3.5-days periods. Interestingly, in Mexico or Peru, the minima are on Sundays-Tuesdays, while in US (Fig. 3 (top)) or Brazil (Fig. 3 (bottom)) they have the pattern like in Europe. It is worth mentioning that North America is currently (May 2020) undergoing suppression of COVID-19 and South America its strong expansion, as expected in the winter-type pattern, which might reappear as a second wave in the Northern Hemisphere.

**Fig. 3.**
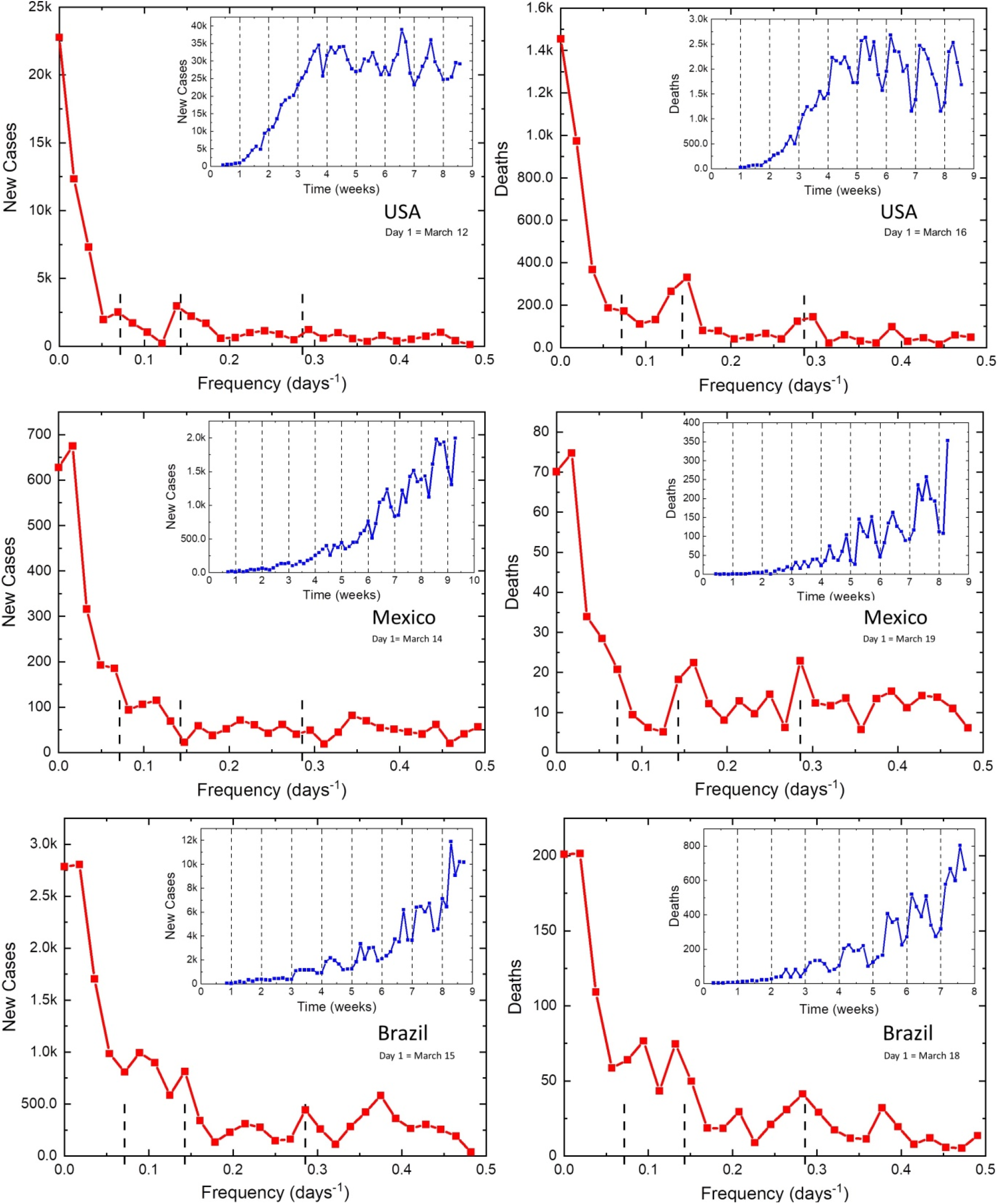
Daily new cases and deaths of COVID-19 reported in USA (bottom), Mexico (middle), and Brazil (bottom).

### Asia

We have also picked two countries from Asia, who provided clear data and were successful in suppressing COVID-19, namely Japan (Fig. 4 (top)) and South Korea (Fig. 4 (bottom)). They both show 7, and 3.5-days periods in infectivity, but rather complex pattern in death rates, perhaps affected by the highly controlled nature of the infectivity.

**Fig. 4.**
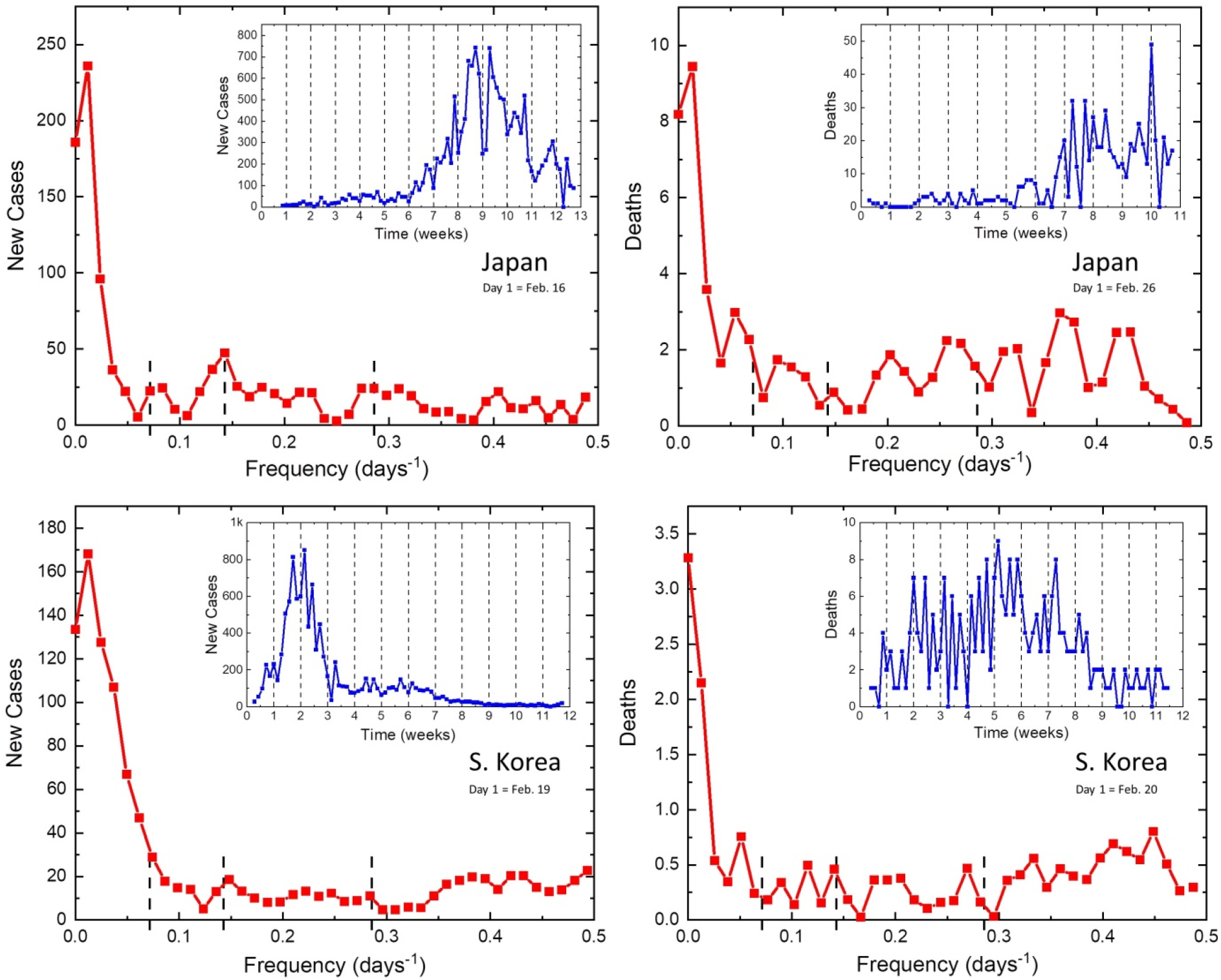
Daily new cases and deaths of COVID-19 reported in Japan (top) and South Korea (bottom).

### World

Finally, we have also provided the analysis in the whole World (Fig. 5), since the collective data became recently available. The results reveal 7, and 3.5-days periods in infectivity and even stronger peaks in death rates. The observed highly regular oscillations of both parameters present in the whole World signifies the globality of these observations and relevance of the obtained data. Despite the fact that the oscillatory dynamics is clearly visible in many countries, some large countries, such as India, do not reveal it.

**Fig. 5.**
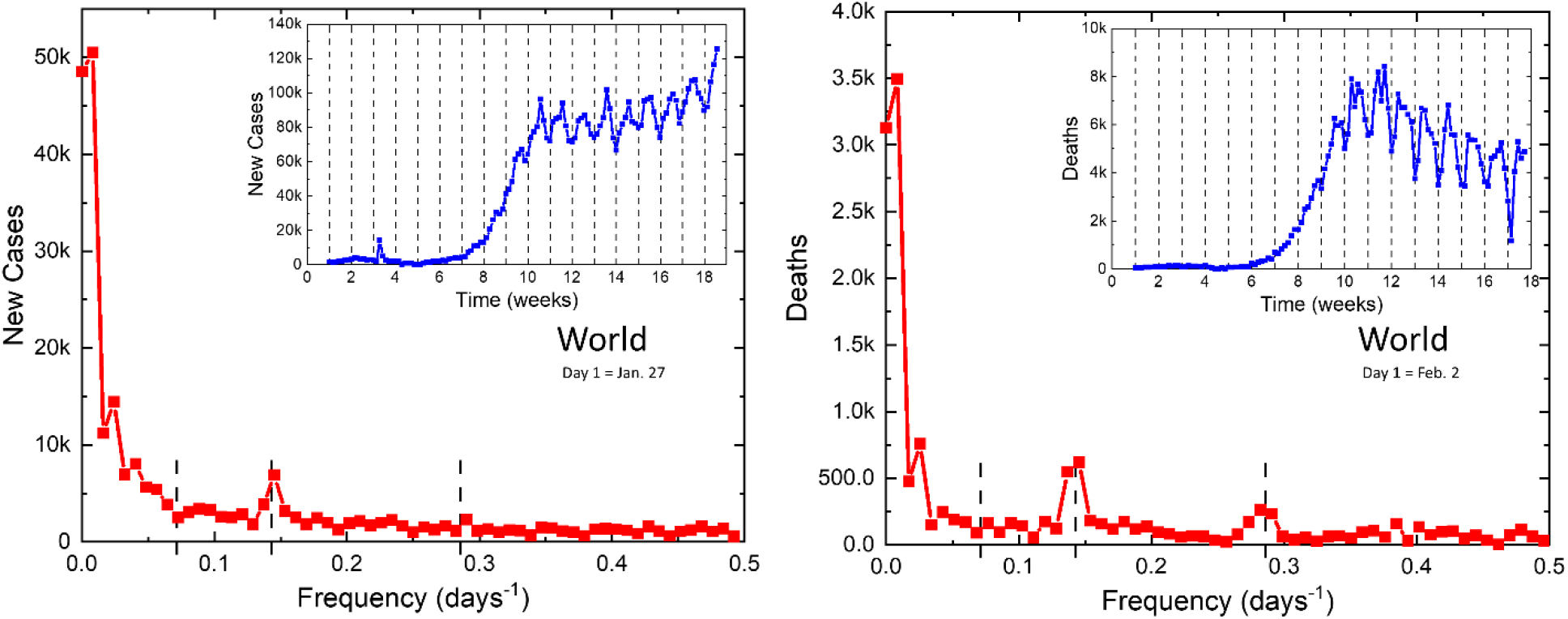
Daily new cases and deaths of COVID-19 reported in the whole World.

## Discussion

Even if we do not know how the circaseptan cyclicity is generated, we could try to set some hypothesis on the underlying mechanisms. We can assume that most of the periodicity observed in the new cases and deaths occurring in COVID-19 does not originate from the sampling stochasticity, but it results from underlying epigenetic modifications^13^ which SARS-HCoV-2 is responsible for. This does not exclude the possibility that epigenetic modifications were triggered by changes of meteorological and pollution variables that exhibit a week-long periodicity^11^. For example, such variables might be human traffic flow outdoor or a poorly working indoor ventilation in hospitals^15^. Finally, we cannot exclude a possibility of the desynchronization of the circadian rhythm. For example, it was observed that rectal temperature and urinary temperature desynchronized in a woman staying approximately three months in isolation in a subterranean cave from a circatrigintan rhytm (i.e., an oscillation with a frequency of 1 cycle in 30 ± 5 days)^16^ and from an internal circasemiseptan (with a frequency of 1 cycle in 3.5 ± 1 days) ^17^

Therefore, we think that it is not a coincidence that the smallest values of the number of new cases and deaths caused by COVID-19 were often optimistically broadcasted on Sundays/Mondays only to be taken again over by the largest values in the coming Fridays/Saturdays. It seems that this 7-days (± 1-day) periodicity in death minima and maxima contradict the hypothesis that patients with serious medical conditions are more likely to die in the hospital if they are admitted on a weekend than if they are admitted on a weekday^14^.

In summary, the described circaseptan (7-days ± 1 day) periodicity pattern, in some cases accompanied by admixture with other infradian cycles, such a hemicircaseptan one, is robust in large and reliably reporting countries representing Europe (Italy; Germany, Sweden, UK), Asia (Japan, South Korea), North (the USA) and South America (Brazil), as well as the whole World. Understanding the relationship between variables associated with the observed periodicities might help with improving health care, better forecasting of the coronavirus infection, stock market expectations about economic growth’s and it might be of interest for providing a clue for the most suitable times at which COVID-19 expected therapies should be administered^18^.

## Materials and Methods

The number of newly detected cases and death rates from COVID-19 in individual countries are publicly available at the following site: https://www.worldometers.info/coronavirus/. The database updating once per day prevents detection of the circadian (24-hour) and of the ultradian periodicities. Since, the time frame of the available data covers mid-February till mid-May, we could not detect periodicities longer than ~15 days (e.g. the lunar one). Also, the data presented in the above-mentioned database were probably somewhat underestimated. The data underestimations result especially due to substantial asymptomatic and pre-symptomatic transmissions that make containment-based interventions, especially those depending on recognition of early symptoms or limited testing, more challenging and potentially infeasible^1^.

For each country, we recorded (for a finite number of days) the daily number of new cases and deaths. Then, using http://lampx.tugraz.at/~hadley/num/ch3/3.3a.php, we performed a Fourier transform of these dependencies and wrote them as follows,

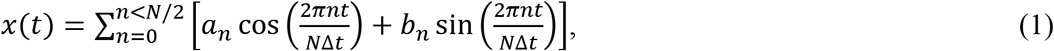

where N is the number of days that we recorded (i.e. the number of data points), Δt = 1 day, 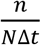 is the frequency, and a_n_, b_n_ are the Fourier coefficients. From these Fourier coefficients, a_n_, b_n_, we calculated the power spectrum, Sxx, and its square root, 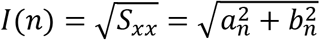, which was depicted and further discussed.

## Data Availability

All data are available upon request.

## Notes

### Competing Interest Statement

The authors have declared no competing interest.

### Funding Statement

No external funding was used.

